# Prognostic value of plasma brain-derived pTau

**DOI:** 10.64898/2026.06.26.26356597

**Authors:** Valentina Ghisays, Marisa N. Denkinger, Alpana Singh, Tainá M. Marques, Michael Malek-Ahmadi, Kendall Van-Keuren Jensen, Hillary D. Protas, Javad Sohankar, Dhruman D. Goradia, Vivek Devadas, Yinghua Chen, Shan Li, Jessica B. Langbaum, Michael W. Weiner, Eric M. Reiman, Yi Su, Nicholas J. Ashton, Alzheimer’s Disease Neuroimaging Initiative

## Abstract

**Background:** Plasma brain-derived pTau217 (BD-pTau217) may provide a Alzheimer’s disease-specific plasma tau measure than total pTau217, but its prognostic value is unclear. We compared BD-pTau217 and total plasma pTau217 for predicting clinical and amyloid PET progression in cognitively unimpaired (CU) ADNI participants.

**Methods:** Plasma NULISAseq biomarkers were measured in 1,427 ADNI participants, including 529 CU individuals. Amyloid PET progression was assessed in baseline CU amyloid-negative participants (Centiloid ≥24.1) with longitudinal PET imaging; clinical progression was assessed in all baseline CU participants. Associations were evaluated using Cox models and time-dependent AUC.

**Results:** BD-pTau217 did not clearly outperform total pTau217 for predicting progression to mild cognitive impairment or dementia. However, among baseline amyloid-negative participants (N=175), BD-pTau217 better predicted amyloid PET positivity at 2.5 years (tdAUC 0.82 vs 0.69; HR=10.54, p=0.00015) and 4 years (tdAUC 0.77 vs 0.64; HR=7.03, p=0.00055).

**Conclusion:** BD-pTau217 improved prediction of near-term amyloid PET progression, with less clear advantage for clinical progression.

**HIGHLIGHTS:**

- BD-pTau217 predicted amyloid PET progression in CU ADNI participants.
- BD-pTau217 outperformed total pTau217 at the CL ≥24.1 threshold.
- BD-pTau217 and total pTau217 performed similarly for clinical progression.
- Prognostic performance varied across amyloid PET thresholds.
- BD-pTau217 may support risk stratification in prevention trials.

**RESEARCH IN CONTEXT:** *Systematic review:* Plasma pTau217 biomarkers show strong cross-sectional associations with amyloid and tau pathology and are increasingly used to support Alzheimer’s disease diagnosis and trial screening. Recent assay designs selectively measuring brain-derived pTau217 may reduce peripheral contributions to total pTau217 measurements, but their longitudinal prognostic value relative to total pTau217 remains less well established.

*Interpretation:* In cognitively unimpaired ADNI participants, plasma BD-pTau217 showed stronger prognostic performance than total pTau217 for progression from amyloid PET negative to amyloid PET positive status, particularly at the primary CL ≥24.1 threshold. In contrast, BD-pTau217 and total pTau217 showed more similar performance for predicting clinical progression to MCI or dementia.

*Future directions:* These findings support further evaluation of BD-pTau217 for biological risk stratification and prevention trial enrichment. Validation is needed in more diverse cohorts, longer follow-up intervals, and across clinical-grade assay platforms.

## 1. INTRODUCTION

The ability to estimate the presence of Alzheimer’s disease (AD) pathology through blood-based measures represents a major advancement in both AD research and, increasingly, clinical care. In symptomatic individuals, elevated levels of phosphorylated tau (pTau217) in the blood strongly suggest that the presenting symptomatology is attributable to AD, as a strong indicator of neuritic amyloid plaque pathology. Conversely, low levels indicate that AD pathology is unlikely to be present at a clinically significant level, and that the observed cognitive deficits may be better explained by an alternative cause.^1^

It is consistently reported that the positive predictive value (PPV) of pTau217 is lower than its negative predictive value (NPV). This difference is partly attributable to lower disease prevalence in certain tested populations (*e.g.,* cognitively unimpaired) but also reflects the fact that elevated pTau217 levels can be occasionally observed due to other reasons, such as chronic kidney disease (CKD) or other medical conditions^2^. In these instances, only more severe cases are likely to elevate pTau217 sufficiently to shift an individual into a different classification group.^3^ It has been shown that in patients with amyotrophic lateral sclerosis (ALS), p-tau levels, including pTau217, are increased in the blood^4,5^ but not cerebrospinal fluid (CSF).^6^ This points to a potentially specific peripheral expression of p-tau—localized, for example, to striated muscle tissue in ALS^4^—but also introduces uncertainty regarding other possible sources of circulating p-tau if such tests are widely adopted in unselected, heterogeneous primary and secondary care populations.

Therefore, to mitigate potential peripheral contributions—whose frequency and impact remain uncertain, the use of brain-derived (BD) pTau217 immunoassays is essential. Tau expressed in the periphery includes a 254–amino acid insert encoded by exon 4a, commonly referred to as “big tau.” Because the sandwich immunoassay design of most pTau217 tests targets epitopes that span regions flanking exon 4a, these assays are not able to distinguish between peripheral pTau217 (if present) and pTau217 released from the central nervous system (CNS) in response to AD pathology. A BD-pTau217 immunoassay can be designed using the same capture antibody targeting phosphorylated threonine 217, but with a detector antibody raised against the junction created when exon 4a is absent. In this configuration, the assay selectively recognizes CNS-type tau and excludes “big tau” isoforms containing the exon 4a insert. To demonstrate this, Janelidze and colleagues compared different plasma pTau217 assay designs in ALS patients and found increases in total pTau217, but not BD-pTau217.^6^ Therefore, with a more specific assay design, enhanced diagnostic performance of BD-pTau217 for AD pathology has been reported.^7–9^ In AD, cross-sectional studies suggest that BD-pTau217 offers modest diagnostic advantages over total plasma pTau217 for identifying cerebral AD pathology.^7^

However, to date, there has not been an evaluation of the prognostic and longitudinal performance of BD-pTau217 (and other BD-pTau assays) compared to total pTau. In this study, we sought to address this knowledge gap by assessing the ability of plasma BD-pTau217 to predict amyloid PET and clinical progression in the Alzheimer’s Disease Neuroimaging Initiative (ADNI) cohort.

## 2. METHODS

### 2.1. Study Participants

Data were obtained from the ADNI cohort. Participants with available plasma NULISAseq CNS 120 panel biomarker measurements were included. Diagnostic classification at baseline followed ADNI criteria for cognitively unimpaired (CU), mild cognitive impairment (MCI), or AD dementia. Demographic characteristics including age, sex, education, and average biomarker values are shown in **Table 1**. For longitudinal analyses, participants were required to have baseline NULISAseq measurement and longitudinal amyloid PET or clinical diagnostic follow-up. Prognostic analyses focused on baseline CU participants: amyloid PET progression analyses were restricted to CU participants without amyloid PET positivity at baseline, whereas clinical progression analyses followed CU participants for progression to MCI or AD dementia. All analyses were performed with publicly available de-identified data from ADNI1/GO/2/3 participants (**Figure 1**).

**Figure 1.**
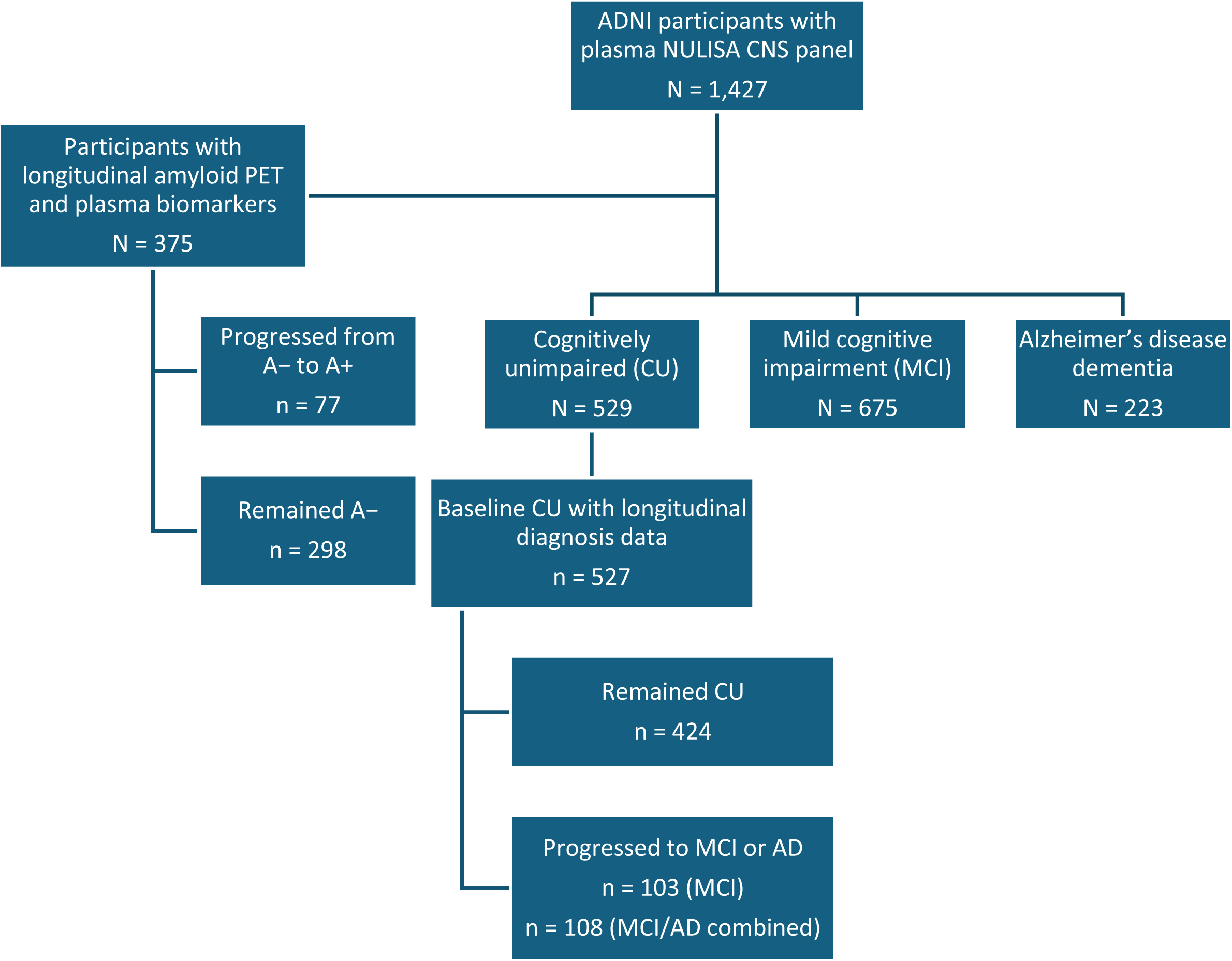
Study design and analytic cohorts. Flow diagram of participants from the Alzheimer’s Disease Neuroimaging Initiative (ADNI) with available plasma NULISAseq CNS panel measurements (N = 1,427). Participants were categorized by baseline clinical diagnosis (cognitively unimpaired [CU], mild cognitive impairment [MCI], or Alzheimer’s disease dementia). Clinical progression analyses included baseline CU participants with longitudinal diagnostic follow-up (n = 527). Amyloid PET analyses were drawn from participants with longitudinal amyloid PET and plasma biomarker data; the primary amyloid PET progression analysis was restricted to baseline CU participants who were amyloid-negative at baseline at the primary CL ≥24.1 threshold and had longitudinal amyloid PET follow-up (n = 175). Subsets were included in longitudinal analyses of (1) amyloid PET progression among participants with baseline amyloid-negative (A−) status and follow-up PET imaging and (2) clinical diagnostic progression among baseline CU participants with longitudinal diagnostic data. Sample sizes differ across analytic cohorts due to availability of longitudinal imaging, baseline amyloid status, clinical diagnosis, and clinical follow-up.

**Table 1.** Baseline demographic and clinical characteristics of the plasma NULISAseq cohort.

|  | CU<br>(N=529) | MCI<br>(N=675) | AD dementia<br>(N=223) | Total<br>(N=1427) | p-value |
| --- | --- | --- | --- | --- | --- |
| Age, years | 72.9 ± 6.3 | 72.4 ± 7.6 | 74.7 ± 8.2 | 73.0 ± 7.3 | <0.001 |
| Female, n (%) | 310 (58.6%) | 276 (40.9%) | 89 (39.9%) | 675 (47.3%) | <0.001 |
| Education, years | 16.7 ± 2.5 | 16.1 ± 2.7 | 15.5 ± 2.9 | 16.2 ± 2.7 | <0.001 |
| Amyloid PET CL | 20.2 ± 37.0 | 46.7 ± 53.1 | 93.9 ± 44.0 | 43.1 ± 52.0 | <0.001 |

Baseline demographic and clinical characteristics of ADNI participants with plasma NULISaseq measurements, overall and by baseline diagnosis group. P-values reflect overall group comparisons across CU, MCI, and AD dementia using ANOVA for continuous variables and chi-square test for sex. Values are mean ± SD unless otherwise indicated.
CU, cognitively unimpaired; MCI, mild cognitive impairment; AD dementia, Alzheimer's disease dementia; CL, Centiloid.

### 2.2. Image analysis

Amyloid PET data ([18F] florbetapir PET scans) from baseline and follow-up were preprocessed with an in-house PET unified pipeline (PUP) ^10–12^ using corresponding baseline T1-weighted structural MRI scans for anatomical labelling in individual space with FreeSurfer v7 (http://surfer.nmr.mgh.harvard.edu/).^13^ Briefly, for semi-quantitative Aβ burden, mean cortical standard uptake value ratios (mcSUVRs) were extracted and averaged across the set of FreeSurfer-defined cortical regions of interest including frontal, parietal, temporal, and precuneus.^10,12^ We converted the mcSUVRs calculated with a cerebellar reference region^14^ to centiloids (CL).^15^

Amyloid PET positivity (A+) was defined using a primary threshold of CL ≥24.1. For cross-sectional analyses, amyloid PET status was determined with the closest visit associated with the NULISAseq measurement and for longitudinal assessments, follow-up scans were used to assess progression from amyloid-negative (A−) to A+. In sensitivity analyses, alternative literature-supported thresholds for A+ were additionally evaluated (Centiloid ≥18 and ≥37).^16,17^ These analyses were performed to assess robustness of biomarker performance across commonly used definitions of A+.

### 2.3. Biomarker measurements

Plasma biomarkers were measured at Michael T. Zuendel Biomarker Laboratory at Banner Sun Health Research Institute on an ARGO HT platform (Alamar Biosciences) using the NULISAseq^TM^ CNS Disease Panel.^18–20^ We use the NULISA Protein Quantification (NPQ) values on the log2 scale with intra-and-inter plate normalization. The highly multiplexed panel measures neurodegenerative disease-related biomarkers in plasma or CSF with ultra-broad dynamic range that can quantify extremely low concentrations of biomarkers like pTau217 in the attomolar range along with highly concentrated proteins from the same 250 μL sample.

Brain-derived and total versions of phosphorylated tau biomarkers were evaluated, with BD-pTau217 designated as the primary biomarker of interest based on prior evidence of strong diagnostic performance. The primary comparison focused on BD-pTau217 versus total pTau217. BD-pTau181 was included as an additional brain-derived phosphorylated tau comparator in the primary amyloid progression table, while other pTau species, GFAP, and pTau/Aβ ratios were evaluated in secondary sensitivity analyses.

### 2.4. Longitudinal outcomes

Longitudinal outcomes included amyloid PET progression and clinical diagnostic progression. For amyloid progression analyses, baseline was defined as the earliest available amyloid PET assessment with a matched baseline plasma NULISAseq measurement. Analyses were restricted to CU participants who were A− at baseline, as defined in Section 2.2, and who had longitudinal amyloid PET follow-up. Participants were followed to progression to A+ status or censored at their last available PET visit if they remained A−. *Time-to-event* was calculated from the baseline A− PET visit to the first A+ PET visit or last follow-up. For sensitivity analyses, amyloid progression was also evaluated using alternative thresholds of CL ≥18 and CL ≥37.^16,17^ Baseline biomarker measurements were matched to the baseline amyloid PET assessment using the closest available plasma sample within a 365-day window.

For clinical progression analyses, baseline diagnosis was defined according to the earliest available longitudinal diagnostic assessment. CU participants were followed for progression to MCI or MCI/AD. Individuals who did not meet progression criteria were censored at their last available diagnostic follow-up. *Time-to-event* was defined as the time interval between baseline plasma biomarker measurement and the first visit at which they progressed.

### 2.5. Statistical analysis

Longitudinal prognostic performance of the plasma biomarkers was assessed using complementary time-to-event and discrimination approaches. Cox proportional hazards (CPH) models were used to estimate hazard ratios (HRs) for amyloid and clinical progression associated with baseline biomarker levels. For binary comparisons, biomarker groups were defined using Youden index-derived cutoffs estimated from receiver operating characteristic (ROC) analyses. These cutoffs were used to define “high” and “low” biomarker groups for Kaplan-Meier (KM), log-rank, and Cox model comparisons.

KM curves were used to visualize risk separation between biomarker-defined high and low groups over time. For amyloid progression analyses, Cox models, log-rank tests, C-indices, and tdAUCs were evaluated at prespecified 2.5- and 4-year horizons by censoring follow-up at each horizon. For clinical progression analyses, Cox models, log-rank tests, C-indices, and full-follow-up AUCs were evaluated across the full available follow-up, while tdAUCs were calculated at 2.5 and 4 years to summarize short-term discrimination. Analyses focused on relative discrimination and ranking performance rather than absolute risk prediction.

We compared BD and total pTau biomarkers using paired analyses where applicable. Statistical significance was evaluated using two-sided tests. In secondary adjusted Cox models, continuous biomarker values were standardized, and models were adjusted for age, sex, and APOE ε4 status to evaluate whether associations between baseline BD-pTau217 and amyloid PET progression were independent of these demographic and genetic risk factors. Analyses were performed in R (version 4.4.3) using the survival, riskRegression, timeROC, and pROC packages.

## 3. RESULTS

### 3.1. Baseline characteristics

We included 1,427 participants with plasma NULISAseq biomarker measurements with a mean age 73.0 (SD ±7.3 years). Baseline age, sex distribution, education, and amyloid PET CL values differed across diagnostic groups (all p < 0.001; **Table 1**). AD participants were older on average and had higher amyloid PET CL values than CU and MCI participants, while the CU group had a higher proportion of women. Prognostic analyses focused on baseline CU participants, including CU participants with longitudinal clinical follow-up for clinical progression analyses and CU participants who were A− at baseline with longitudinal amyloid PET follow-up for amyloid PET progression analyses.

### 3.2. Amyloid PET progression

Among CU participants with available baseline amyloid PET, 186 were A− and 76 were A+ at the primary CL ≥24.1 threshold. Baseline plasma BD-pTau217 levels were higher in CU A+ than CU A− participants (median 11.9 [IQR 11.6–12.3] vs 11.0 [10.7–11.2] NPQ), as were total pTau217 levels (median 11.8 [11.3–12.1] vs 10.6 [10.3–11.0] NPQ). For longitudinal amyloid progression analyses, 175 baseline CU A− participants with longitudinal amyloid PET follow-up were included. At the primary amyloid positivity threshold (CL ≥24.1), BD-pTau217 showed the strongest prognostic performance, whereas total pTau217 and BD-pTau181 showed weaker and non-significant associations (**Table 2**). BD-pTau217 showed the highest time-dependent discrimination across follow-up, with prespecified 2.5- and 4-year estimates reported in Table 2 and the broader tdAUC trajectory is shown in Figure 2A. This pattern was also apparent in the KM analyses, in which BD-pTau217 showed clearer risk separation than total pTau217 over follow-up (**Figure 2B-C**). At this primary threshold, most ratio measures and alternative pTau species, including BD-pTau231 and pTau231-based measures, showed weaker or non-significant associations, supporting BD-pTau217 as the strongest overall predictor.

**Figure 2.**
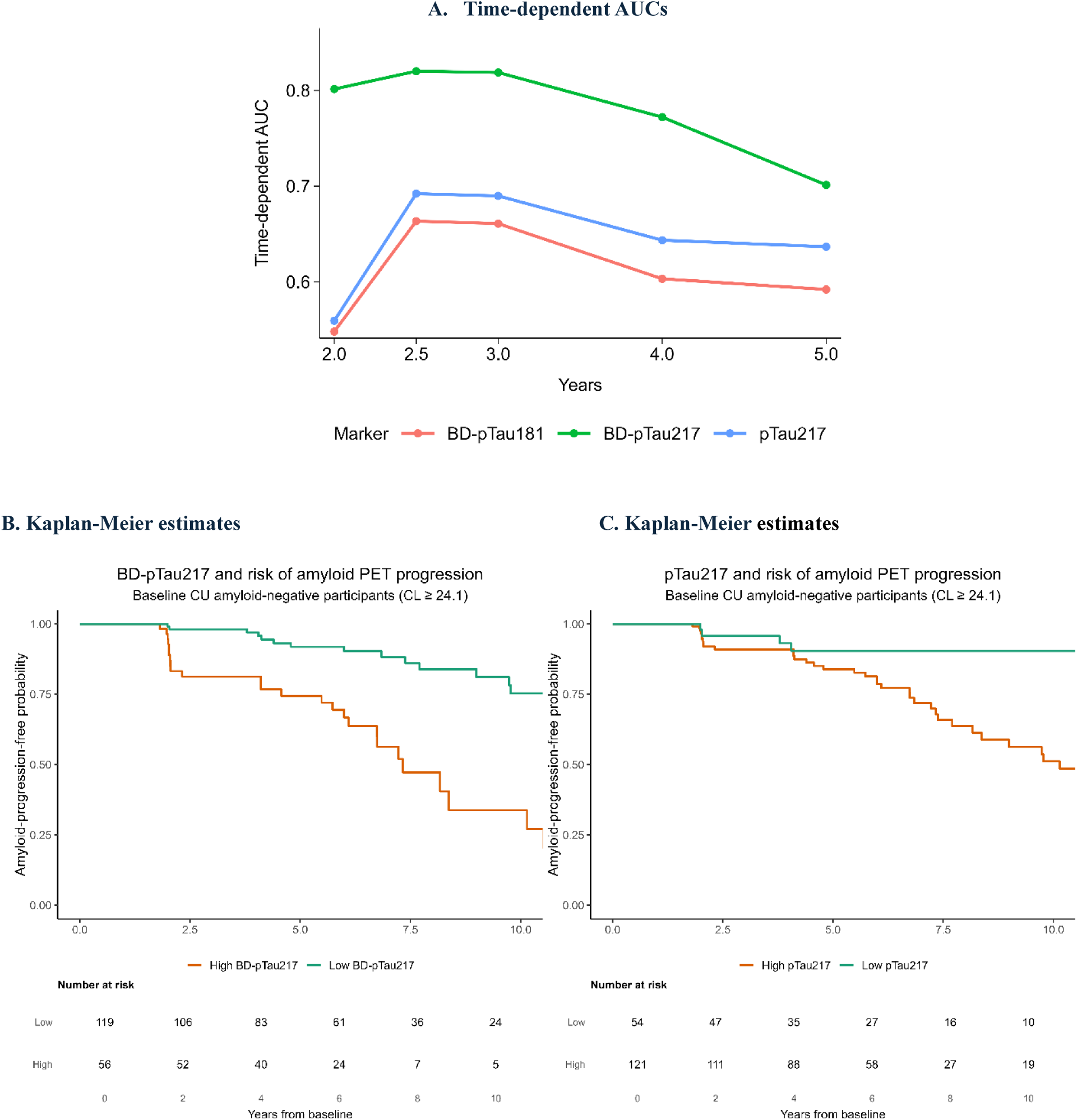
Prognostic performance of plasma pTau biomarkers for amyloid PET progression. (A) Time-dependent AUC trajectories for prediction of amyloid PET progression (A− to A+) among baseline cognitively unimpaired (CU) participants at the primary amyloid positivity threshold (CL≥24.1). Table 2 reports the prespecified 2.5- and 4-year estimates. BD-pTau217 demonstrated consistently higher discrimination than total pTau217 and BD-pTau181. (B-C) Kaplan-Meier (KM) estimates of amyloid-progression-free probability according to high versus low baseline BD-pTau217 (B) and total pTau217 (C), defined using Youden index cutoffs. Curves reflect cumulative risk separation over the full available follow-up period among CU participants who were A− at baseline (CL ≥24.1).

**Table 2.** Prognostic performance of plasma biomarkers for amyloid PET progression (A− to A+) in baseline. cognitively unimpaired participants at the primary amyloid positivity threshold (Centiloid ≥ 24.1).

| Marker | tdAUC<br>(2.5y) | HR (95% CI)<br>high vs low | Log-rank<br>p | C-index | tdAUC<br>(4y) | HR (95% CI)<br>high vs low | Log-rank<br>p | C-index |
| --- | --- | --- | --- | --- | --- | --- | --- | --- |
| BD-pTau217 | 0.82 | 10.54 (2.31-48.10) | 0.00015 | 0.76 | 0.77 | 7.03 (1.94-25.56) | 0.00055 | 0.73 |
| pTau217 | 0.69 | 2.15 (0.47-9.79) | 0.31267 | 0.57 | 0.64 | 1.42 (0.39-5.17) | 0.58945 | 0.54 |
| BD-pTau181 | 0.66 | 2.33 (0.63-8.60) | 0.19180 | 0.59 | 0.60 | 1.75 (0.54-5.69) | 0.34484 | 0.57 |

Discrimination is summarized using time-dependent area under the curve (tdAUC) at prespecified 2.5- and 4-year horizons. Hazard ratios (HRs) and C-indices were derived from Cox proportional hazards models comparing high versus low biomarker groups within each follow-up horizon. Log-rank p-values compare Kaplan-Meier (KM) curves for the same high versus low biomarker groups. High versus low biomarker groups were defined using Youden index cutoffs derived from overall amyloid progression status across the full follow-up period.

At 2.5 years (N = 175; converters = 12), elevated baseline BD-pTau217 was strongly associated with amyloid PET progression (HR 10.54, 95% CI 2.3–48.1; log-rank p = 0.00015; tdAUC 0.82). In contrast, total pTau217 and BD-pTau181 were not significantly associated with progression and showed substantially lower discrimination. At 4 years, BD-pTau217 remained the strongest predictor (HR 7.03, 95% CI 1.9–25.6; log-rank p = 0.00055; tdAUC 0.77), while total pTau217 and BD-pTau181 again demonstrated weak and non-significant associations (**Table 2**). These results suggest that prognostic performance at the primary threshold was strongest for BD-pTau217 rather than reflecting a uniform signal across pTau measures.

Across sensitivity analyses, biomarker performance varied according to the amyloid positivity threshold used (**Supplementary Tables S1–S2**). At the lower threshold (CL ≥18), discrimination was generally weaker and less consistent across single biomarkers, although selected pTau/Aβ42 ratio measures, including BD-pTau217/Aβ42 and pTau217/Aβ42, showed stronger associations with amyloid progression compared with their corresponding single-marker measures.

At the higher threshold (CL ≥37), several biomarkers and ratio measures demonstrated improved prognostic performance, including total pTau217, BD-pTau181, BD-pTau231, and corresponding pTau/Aβ42 ratio measures, consistent with increasing signal at more advanced stages of amyloid burden. Notably, at both CL ≥18 and CL ≥37, estimates at the 2.5- and 4-year horizons were identical because all observed amyloid conversion events occurred within the first 2.5 years of follow-up (e.g., CL ≥18: 14 events by 2.5 years and 4 years; CL ≥37: 13 events by 2.5 years and 4 years), whereas at the primary threshold (CL ≥24.1), additional events occurring after 2.5 years resulted in differing estimates between time horizons. Overall, BD-pTau217 demonstrated the most robust and consistent prognostic performance across thresholds, remaining the strongest predictor at the clinically relevant CL ≥24.1 threshold.

In secondary adjusted Cox models, baseline BD-pTau217 remained associated with amyloid PET progression after adjustment for age, sex, and APOE ε4 status at both 2.5 years (HR 1.45 per SD, 95% CI 1.08-1.95; p = 0.014) and 4 years (HR 1.40 per SD, 95% CI 1.04-1.89; p = 0.027).

### 3.3. Clinical progression

Among the 527 baseline CU participants included, 108 progressed to MCI or AD dementia during a median follow-up of 5.9 years (IQR 3.0-7.2). When restricting progression to MCI only, 103 participants progressed and 424 remained CU.

Participants with high total pTau217 levels had a 2.8-fold increased risk of progression compared with those in the low group (HR 2.8, 95% CI 1.86-4.07; log-rank p<0.001), with progression observed in 31.8% of the high group versus 12.8% of the low group. BD-pTau217 showed comparable risk estimates (HR 2.7, 95% CI 1.83-3.89; high group 34.8% vs low group 14.5% progressed). Similar risk estimates were observed for BD-pTau181 (HR 2.6, 95% CI 1.73-3.86). Full-follow-up AUCs were similar across markers (0.65-0.66), and concordance indices ranged from 0.61 to 0.63. Time-dependent discrimination at 2.5 and 4 years was also modest and overlapped across markers (tdAUCs approximately 0.59-0.61) (**Table 3**, **Figure 3**). KM analyses showed separation for both BD-pTau217 and total pTau217, but overall differences between markers were smaller than for amyloid PET progression (**Figure 3B-C**).

**Figure 3.**
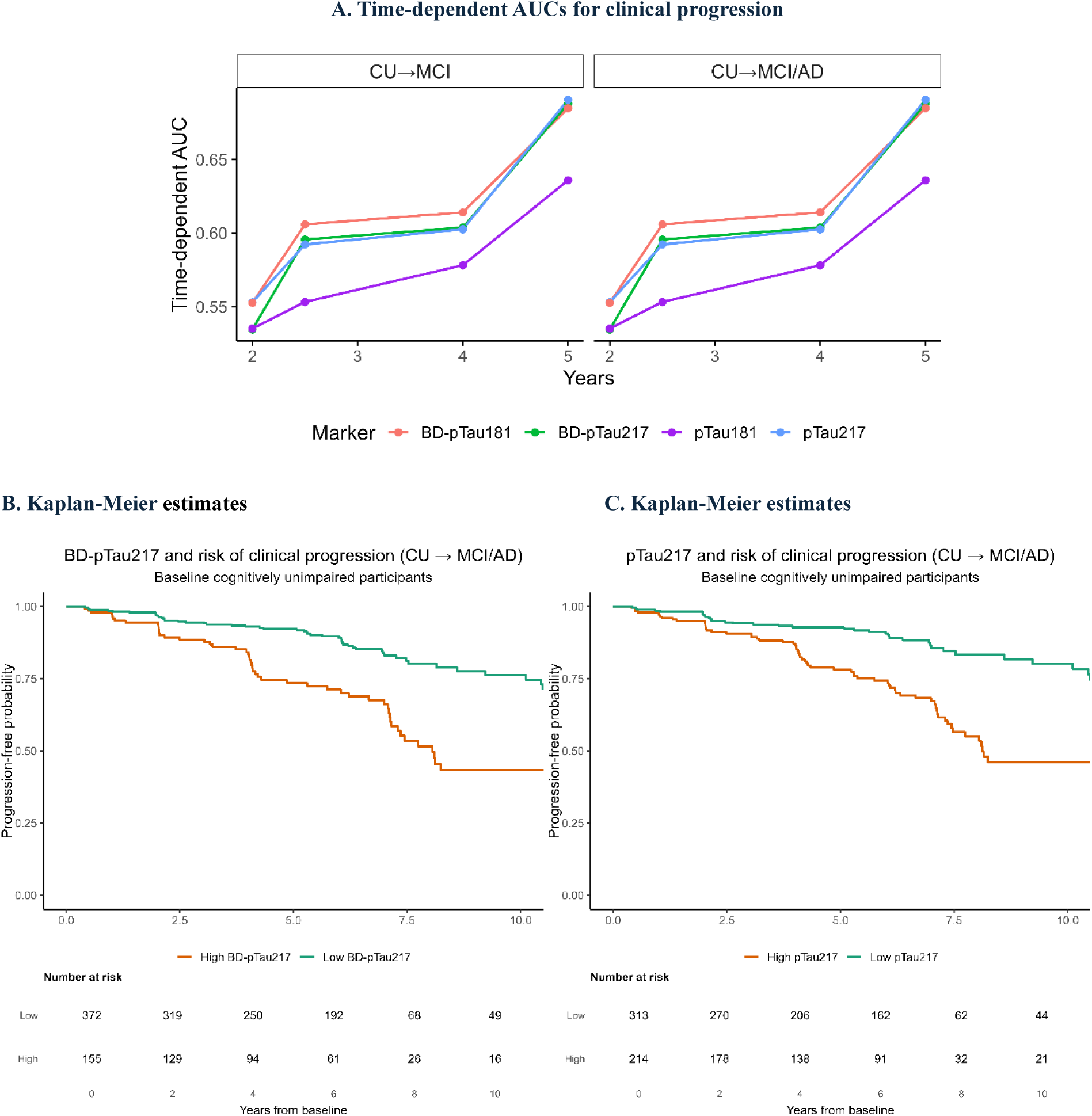
Prognostic performance of plasma pTau biomarkers for clinical progression. (A) Time-dependent discrimination (tdAUC) for prediction of progression from cognitively unimpaired status to mild cognitive impairment (CU→MCI) and to CU→MCI/AD across prespecified follow-up horizons using baseline plasma biomarkers. Discrimination was modest and similar across biomarkers. (B) Kaplan-Meier (KM) estimates of progression-free probability according to baseline BD-pTau217 level dichotomized using a Youden index cutoff among baseline CU participants. (C) KM estimates of progression-free probability according to baseline total pTau217 level dichotomized using a Youden index cutoff among baseline CU participants. Higher biomarker levels were associated with greater cumulative risk of clinical progression over the full available follow-up period.

**Table 3.** Prognostic performance of plasma pTau biomarkers for clinical progression from cognitively unimpaired to MCI or AD dementia.

| Outcome | Marker | tdAUC 2.5y | tdAUC 4y | AUC full follow-up | HR (95% CI) high vs low | Log-rank p | C-index |
| --- | --- | --- | --- | --- | --- | --- | --- |
| CU→MCI/AD | pTau217 | 0.59 | 0.60 | 0.66 | 2.8 (1.86-4.07) | <0.001 | 0.62 |
| CU→MCI/AD | BD-pTau217 | 0.60 | 0.60 | 0.66 | 2.7 (1.83-3.89) | <0.001 | 0.61 |
| CU→MCI/AD | BD-pTau181 | 0.61 | 0.61 | 0.65 | 2.6 (1.73-3.86) | <0.001 | 0.63 |
| CU→MCI | pTau217 | 0.59 | 0.60 | 0.66 | 2.8 (1.86-4.15) | <0.001 | 0.62 |
| CU→MCI | BD-pTau217 | 0.60 | 0.60 | 0.65 | 2.6 (1.78-3.86) | <0.001 | 0.61 |
| CU→MCI | BD-pTau181 | 0.61 | 0.61 | 0.65 | 2.6 (1.74-3.97) | <0.001 | 0.63 |

Time-dependent areas under the curve (tdAUCs) summarize discrimination at prespecified 2.5- and 4-year horizons. Full-follow-up AUC summarizes continuous biomarker discrimination for overall clinical progression status across available follow-up. Hazard ratios (HRs), log-rank *p*-values, and C-indices are derived from full-follow-up Cox and Kaplan-Meier (KM) analyses comparing high versus low biomarker groups. High versus low groups were defined using Youden index cutoffs derived from overall clinical progression status. CU→MCI and CU→MCI/AD yielded similar tdAUC estimates at the 2.5- and 4-year horizons because nearly all early clinical progression events captured by these horizons were transitions from CU to MCI.

Results were similar when we restricted progression to MCI alone, where high total pTau217 was associated with a 2.8-fold increased risk of progression (95% CI 1.86-4.15), with progression rates of 30.4% in the high group versus 12.1% in the low group. BD-pTau217 again had comparable performance (HR 2.6, 95% CI 1.78-3.86; progression rates 32.9% versus 14.0% in high and low groups, respectively), again with overlapping discrimination metrics across biomarkers (**Table 3**). Across models, discrimination for clinical progression was modest but consistent.

## 4. DISCUSSION

In this study, we evaluated the prognostic performance of brain-derived and total plasma pTau biomarkers for amyloid PET and clinical progression in cognitively unimpaired ADNI participants with plasma NULISAseq measurements. We found that BD-pTau217 demonstrated stronger prognostic value for amyloid PET progression, particularly at the primary A+ threshold of CL ≥24.1. At this threshold, BD-pTau217 was the only primary marker to show strong and statistically significant prediction at both 2.5-and 4-year progression, whereas total pTau217 and BD-pTau181 showed weaker and non-significant associations. Sensitivity analyses across CL

≥18 and CL ≥37 further suggested that biomarker performance varies by amyloid threshold, with broader pTau and pTau/Aβ42 ratio^21^ signals emerging most clearly with alternative thresholds.

In contrast to findings for amyloid PET progression, differences between brain-derived and total plasma pTau measures for prediction of clinical progression were modest, with both assay designs showing similar discrimination for progression from cognitively unimpaired status to MCI or dementia. Exploratory analyses further indicated that incorporation of plasma Aβ42 into biomarker ratios^21^ did not materially improve prediction beyond pTau217 alone for clinical outcomes. These findings suggest that while plasma pTau217 measures are associated with future clinical change, improvements in biological specificity achieved through brain-derived measurements appear to have greater relevance for predicting underlying pathological progression than for short-term clinical transitions.

Prior studies of plasma pTau biomarkers have largely focused on cross-sectional discrimination of amyloid and tau pathology, consistently demonstrating better performance of plasma pTau217 to discriminate between the presence or absence of amyloid plaques and the diagnosis of AD compared with blood biomarkers.^6,19,22,23^ Recent work has additionally suggested that selectively measuring brain-derived fractions of plasma pTau may improve diagnostic performance.^7–9^ Our results extend these observations by demonstrating that BD-pTau217 provides stronger prognostic information for future amyloid accumulation than total plasma pTau217, particularly at the clinically relevant CL ≥24.1 threshold. Although brain-derived and total pTau217 captures overlapping biological signal, the stronger and more consistent discrimination observed for BD-pTau217 suggests that isolating brain-origin tau species may enhance sensitivity to active amyloid-related neurobiological change by reducing non-specific signal for the periphery.

In analyses focused on amyloid PET progression, the relative performance of biomarkers depended on the amyloid positivity threshold used. At the primary CL ≥24.1 threshold, a common range used in clinical trial design,^16,17^ BD-pTau217 clearly outperformed total pTau217 and BD-pTau181, which showed weak and non-significant associations with progression. At the lower CL ≥18 threshold, single-marker discrimination was weaker and less consistent, while selected pTau/Aβ42 ratio measures^21,23^, including BD-pTau217/Aβ42 and pTau217/Aβ42, showed stronger associations than their corresponding single-marker measures.^24,25^ This supports the notion that Aβ peptides in plasma may play an important role in the earliest phase of cerebral amyloid accumulation, despite challenges in robustness.^26^ At the higher CL ≥37 threshold, a broader set of biomarkers became informative, including total pTau217, BD-pTau181, BD-pTau231, and several pTau/Aβ42 ratio measures. This threshold-dependent pattern suggests that different plasma biomarker combinations may be informative at different stages of amyloid accumulation, while BD-pTau217 remains the most robust single-marker predictor at the primary clinically relevant threshold.

Across clinical endpoints, discrimination for progression from CU to MCI or MCI/AD was modest and largely overlapping across biomarkers. BD-pTau217, total pTau217, and BD-pTau181 showed similar risk estimates and concordance values (HRs, c-indices, and tdAUC), indicating that brain-derived measurements did not clearly improve prediction of clinical progression. In contrast to amyloid PET progression, where BD-pTau217 showed more pronounced advantage at the primary amyloid threshold, differences between brain-derived and total pTau measures were small for clinical outcomes. The weaker separation for clinical outcomes is consistent with clinical progression being influenced by multiple factors beyond amyloid and plasma tau burden, including downstream neurodegeneration, vascular factors, comorbid pathology, and individual resilience.

The stronger prognostic performance observed for amyloid PET progression compared with clinical progression likely reflects the closer biological relationship between plasma pTau biomarkers and underlying AD pathology than with downstream clinical symptoms. Cognitive decline results from a combination of amyloid and tau pathology but also neurodegeneration, vascular factors, comorbid pathology (e.g., TDP-43), and individual resilience, making clinical progression inherently more heterogeneous and difficult to predict. In contrast, progression from A− to A+ status more directly reflects ongoing AD pathological processes, which plasma pTau biomarkers are thought to capture. These findings suggest that plasma pTau measurements, particularly BD-pTau217, may be valuable for identifying cognitively unimpaired, A− individuals at elevated risk of near-term amyloid progression, including those who may be candidates for primary prevention trials aimed at preventing progression of AD pathology.

Several considerations should be noted for the interpretation of these findings. First, plasma biomarkers were measured using the NULISAseq research platform, which provides highly sensitive, multiplexed relative quantification but differs from emerging *in-vitro* diagnostic platforms intended for routine clinical use. Therefore, the absolute performance metrics in this study may not directly translate to the clinical assays, underscoring the need to further validate across independent cohorts and assay platforms.^7,9^ Second, clinical progression endpoints are influenced by multiple pathological and non-pathological factors beyond amyloid and tau, which likely contributes to the moderate discrimination observed for the clinical outcomes. Third, the number of amyloid progression events was limited, particularly in horizon-specific and threshold-specific analyses, which may reduce the precision of some estimates. Lastly, external validation datasets will be important to determine biomarker performance across populations, assay platforms, and follow-up intervals.

To summarize, plasma BD-pTau217 has shown an added advantage in cross-sectional prediction of AD pathology,^7–9^ however, in this study, we demonstrated that BD-pTau217 has strongest and most consistent prognostic performance for amyloid PET progression among cognitively unimpaired participants. At the primary >24.1 CL threshold, BD-pTau217 clearly outperformed both pTau217 and BD-pTau181, whereas differences between brain-derived and total tau measures were smaller for predicting clinical progression. Diagnostic sensitivity analyses indicated that biomarker performance varied across amyloid positivity thresholds, with selected pTau/Aβ42 ratio measures and additional pTau species becoming more informative at alternative cut points. Collectively, these findings support the use of BD-pTau217 for risk stratification in prevention trials targeting the earliest detectable stages of Alzheimer’s disease. Future studies should evaluate its performance in more diverse populations, over longer follow-up periods, and across multiple assay platforms to facilitate clinical implementation.

### Consent Statement

Data used in this study were obtained from the Alzheimer’s Disease Neuroimaging Initiative (ADNI). Written informed consent was obtained from all ADNI participants or their authorized representatives at each participating site, and ADNI protocols were approved by the institutional review boards of all participating institutions. The present study used de-identified ADNI data.

## Data Availability

Data used in the preparation of this article were obtained from the Alzheimers Disease Neuroimaging Initiative (ADNI) database (adni.loni.usc.edu). The ADNI was launched in 2003 as a public-private partnership, led by Principal Investigator Michael W. Weiner, MD. The primary goal of ADNI has been to test whether serial magnetic resonance imaging (MRI), positron emission tomography (PET), other biological markers, and clinical and neuropsychological assessment can be combined to measure the progression of mild cognitive impairment (MCI) and early Alzheimers disease (AD). For up-to-date information, see www.adni-info.org

http://adni.loni.usc.edu/

## Acknowledgments

Data collection and sharing for the Alzheimer’s Disease Neuroimaging Initiative (ADNI) is funded by the National Institute on Aging (National Institutes of Health Grant U19 AG024904). The grantee organization is the Northern California Institute for Research and Education. In the past, ADNI has also received funding from the National Institute of Biomedical Imaging and Bioengineering, the Canadian Institutes of Health Research, and private sector contributions through the Foundation for the National Institutes of Health (FNIH) including generous contributions from the following: AbbVie, Alzheimer’s Association; Alzheimer’s Drug Discovery Foundation; Araclon Biotech; BioClinica, Inc.; Biogen; Bristol-Myers Squibb Company; CereSpir, Inc.; Cogstate; Eisai Inc.; Elan Pharmaceuticals, Inc.; Eli Lilly and Company; EuroImmun; F. Hoffmann-La Roche Ltd and its affiliated company Genentech, Inc.; Fujirebio; GE Healthcare; IXICO Ltd.; Janssen Alzheimer Immunotherapy Research & Development, LLC.; Johnson & Johnson Pharmaceutical Research &Development LLC.; Lumosity; Lundbeck; Merck & Co., Inc.; Meso Scale Diagnostics, LLC.; NeuroRx Research; Neurotrack Technologies; Novartis Pharmaceuticals Corporation; Pfizer Inc.; Piramal Imaging; Servier; Takeda Pharmaceutical Company; and Transition Therapeutics.

