## Supplementary Tables S1 - S2 for "Prognostic value of plasma brain-derived pTau"

Supplementary Table S1. Sensitivity analyses of plasma biomarker prediction of amyloid PET progression across alternative Centiloid thresholds

| **Marker** | **CL**  **threshold** | **tdAUC (95% CI) 2.5y** | **HR (95% CI)** | **Log-rank p** | **C-index** | **tdAUC (95% CI) 4y** | **HR (95% CI)** | **Log-rank p** | **C-index** |
| --- | --- | --- | --- | --- | --- | --- | --- | --- | --- |
| BD-pTau217 | 18 | 0.68 (0.51-0.85) | 2.03 (0.57-7.27) | 0.27 | 0.57 | 0.68 (0.51-0.85) | 2.03 (0.57-7.27) | 0.27 | 0.57 |
| pTau217 | 18 | 0.69 (0.54-0.85) | 2.53 (0.57-11.32) | 0.21 | 0.57 | 0.69 (0.53-0.84) | 2.53 (0.57-11.32) | 0.21 | 0.57 |
| BD-pTau181 | 18 | 0.58 (0.38-0.78) | 1.42 (0.48-4.23) | 0.53 | 0.53 | 0.57 (0.37-0.78) | 1.42 (0.48-4.23) | 0.53 | 0.53 |
| pTau181 | 18 | 0.60 (0.44-0.77) | 2.13 (0.67-6.79) | 0.19 | 0.58 | 0.60 (0.43-0.76) | 2.13 (0.67-6.79) | 0.19 | 0.58 |
| BD-pTau231 | 18 | 0.67 (0.52-0.82) | 5.56 (0.73-42.54) | 0.06 | 0.61 | 0.66 (0.51-0.81) | 5.56 (0.73-42.54) | 0.06 | 0.61 |
| pTau231 | 18 | 0.69 (0.53-0.85) | 4.36 (1.22-15.63) | 0.01 | 0.66 | 0.68 (0.52-0.84) | 4.36 (1.22-15.63) | 0.01 | 0.66 |
| GFAP | 18 | 0.56 (0.41-0.71) | 1.95 (0.65-5.83) | 0.22 | 0.57 | 0.55 (0.40-0.71) | 1.95 (0.65-5.83) | 0.22 | 0.57 |
| BD-pTau217 | 24.1 | 0.82 (0.72-0.92) | 10.54 (2.31-48.10) | 0.00 | 0.76 | 0.77 (0.65-0.90) | 7.03 (1.94-25.56) | 0.00 | 0.73 |
| pTau217 | 24.1 | 0.69 (0.53-0.85) | 2.15 (0.47-9.79) | 0.31 | 0.57 | 0.64 (0.47-0.81) | 1.42 (0.39-5.17) | 0.59 | 0.54 |
| BD-pTau181 | 24.1 | 0.66 (0.47-0.85) | 2.33 (0.63-8.60) | 0.19 | 0.59 | 0.60 (0.40-0.80) | 1.75 (0.54-5.69) | 0.34 | 0.57 |
| pTau181 | 24.1 | 0.55 (0.37-0.72) | 1.77 (0.56-5.58) | 0.32 | 0.57 | 0.50 (0.32-0.68) | 1.55 (0.51-4.72) | 0.44 | 0.55 |
| BD-pTau231 | 24.1 | 0.64 (0.47-0.81) | 3.48 (1.05-11.56) | 0.03 | 0.65 | 0.60 (0.44-0.77) | 2.78 (0.91-8.51) | 0.06 | 0.63 |
| pTau231 | 24.1 | 0.55 (0.37-0.72) | 1.87 (0.59-5.90) | 0.28 | 0.58 | 0.50 (0.32-0.67) | 1.55 (0.52-4.63) | 0.42 | 0.56 |
| GFAP | 24.1 | 0.66 (0.53-0.79) | 3.37 (0.91-12.44) | 0.05 | 0.64 | 0.68 (0.55-0.80) | 3.74 (1.03-13.61) | 0.03 | 0.65 |
| BD-pTau217 | 37 | 0.84 (0.75-0.93) | 10.84 (2.98-39.40) | 0.00 | 0.77 | 0.84 (0.75-0.93) | 10.84 (2.98-39.40) | 0.00 | 0.77 |
| pTau217 | 37 | 0.76 (0.63-0.89) | 8.93 (2.46-32.45) | 0.00 | 0.75 | 0.76 (0.62-0.89) | 8.93 (2.46-32.45) | 0.00 | 0.75 |
| BD-pTau181 | 37 | 0.74 (0.58-0.91) | 4.64 (1.28-16.85) | 0.01 | 0.68 | 0.74 (0.57-0.90) | 4.64 (1.28-16.85) | 0.01 | 0.68 |
| pTau181 | 37 | 0.60 (0.44-0.75) | 2.80 (0.94-8.33) | 0.05 | 0.62 | 0.59 (0.44-0.75) | 2.80 (0.94-8.33) | 0.05 | 0.62 |
| BD-pTau231 | 37 | 0.70 (0.52-0.88) | 7.31 (2.01-26.60) | 0.00 | 0.73 | 0.69 (0.51-0.88) | 7.31 (2.01-26.60) | 0.00 | 0.73 |
| pTau231 | 37 | 0.64 (0.47-0.80) | 4.21 (1.16-15.31) | 0.02 | 0.67 | 0.63 (0.46-0.80) | 4.21 (1.16-15.31) | 0.02 | 0.67 |
| GFAP | 37 | 0.70 (0.57-0.82) | 3.93 (1.32-11.70) | 0.01 | 0.66 | 0.69 (0.57-0.82) | 3.93 (1.32-11.70) | 0.01 | 0.66 |
| Sensitivity analyses evaluating prognostic performance of individual plasma biomarkers for prediction of amyloid PET progression (A− to A+) among baseline cognitively unimpaired participants using alternative amyloid positivity thresholds (Centiloid ≥18, ≥24.1, and ≥37). Discrimination is summarized using time-dependent area under the curve (tdAUC) at prespecified 2.5- and 4-year horizons. Hazard ratios (HRs) and C-indices were derived from Cox proportional hazards models comparing high versus low biomarker groups within each horizon. Log-rank p-values compare Kaplan-Meier curves for the same high versus low biomarker groups. HRs compare high versus low biomarker groups defined by Youden cutoffs within each threshold. CL indicates Centiloid. | | | | | | | | | |

Supplementary Table S2. Prediction of amyloid PET progression using plasma pTau/Aβ42 ratio measures and additional pTau species

| **Marker** | **CL**  **threshold** | **tdAUC (95% CI)**  **2.5y** | **HR (95% CI)** | **Log-rank p** | **C-index** | **tdAUC (95% CI)**  **4y** | **HR (95% CI)** | **Log-rank p** | **C-index** |
| --- | --- | --- | --- | --- | --- | --- | --- | --- | --- |
| BD-pTau217/Aβ42 | 18 | 0.71 (0.58-0.85) | 5.65 (1.77–18.01) | 0.00 | 0.70 | 0.71 (0.57-0.84) | 5.65 (1.77–18.01) | 0.00 | 0.70 |
| pTau217/Aβ42 | 18 | 0.72 (0.57-0.87) | 4.29 (1.20–15.39) | 0.01 | 0.66 | 0.71 (0.57-0.86) | 4.29 (1.20–15.39) | 0.01 | 0.66 |
| BD-pTau181/Aβ42 | 18 | 0.64 (0.46-0.82) | 3.12 (1.05–9.32) | 0.03 | 0.63 | 0.63 (0.46-0.81) | 3.12 (1.05–9.32) | 0.03 | 0.63 |
| pTau181/Aβ42 | 18 | 0.62 (0.44-0.79) | 2.51 (0.70–9.01) | 0.14 | 0.59 | 0.61 (0.44-0.79) | 2.51 (0.70–9.01) | 0.14 | 0.59 |
| BD-pTau231/Aβ42 | 18 | 0.67 (0.52-0.81) | 5.39 (0.71–41.21) | 0.07 | 0.61 | 0.66 (0.52-0.81) | 5.39 (0.71–41.21) | 0.07 | 0.61 |
| pTau231/Aβ42 | 18 | 0.67 (0.51-0.84) | 3.34 (0.93–11.97) | 0.05 | 0.63 | 0.67 (0.50-0.83) | 3.34 (0.93–11.97) | 0.05 | 0.63 |
| Aβ40/Aβ42 | 18 | 0.51 (0.32-0.70) | 3.54 (1.23–10.19) | 0.01 | 0.63 | 0.52 (0.33-0.71) | 3.54 (1.23–10.19) | 0.01 | 0.63 |
| BD-pTau217/Aβ42 | 24.1 | 0.61 (0.47-0.76) | 3.12 (0.99–9.84) | 0.04 | 0.64 | 0.61 (0.47-0.74) | 2.60 (0.87–7.72) | 0.08 | 0.62 |
| pTau217/Aβ42 | 24.1 | 0.62 (0.46-0.78) | 4.00 (1.08–14.79) | 0.02 | 0.66 | 0.59 (0.44-0.75) | 2.99 (0.92–9.72) | 0.06 | 0.64 |
| BD-pTau181/Aβ42 | 24.1 | 0.54 (0.35-0.74) | 2.85 (0.90–8.98) | 0.06 | 0.63 | 0.52 (0.33-0.70) | 2.36 (0.79–7.04) | 0.11 | 0.61 |
| pTau181/Aβ42 | 24.1 | 0.50 (0.34-0.66) | 1.79 (0.39–8.17) | 0.45 | 0.55 | 0.47 (0.32-0.63) | 1.96 (0.43–8.84) | 0.37 | 0.55 |
| BD-pTau231/Aβ42 | 24.1 | 0.43 (0.27-0.58) | 1.08 (0.35–3.34) | 0.90 | 0.51 | 0.45 (0.30-0.59) | 1.25 (0.42–3.73) | 0.68 | 0.52 |
| pTau231/Aβ42 | 24.1 | 0.49 (0.32-0.66) | 1.61 (0.52–4.98) | 0.41 | 0.56 | 0.46 (0.31-0.62) | 1.38 (0.46–4.10) | 0.56 | 0.54 |
| Aβ40/Aβ42 | 24.1 | 0.45 (0.28-0.63) | 0.89 (0.19–4.05) | 0.88 | 0.51 | 0.49 (0.32-0.67) | 1.32 (0.36–4.80) | 0.67 | 0.52 |
| BD-pTau217/Aβ42 | 37 | 0.77 (0.66-0.88) | 6.05 (1.86–19.64) | 0.00 | 0.72 | 0.77 (0.66-0.87) | 6.05 (1.86–19.64) | 0.00 | 0.72 |
| pTau217/Aβ42 | 37 | 0.77 (0.68-0.87) | 14.69 (1.91–113.00) | 0.00 | 0.74 | 0.77 (0.67-0.87) | 14.69 (1.91–113.00) | 0.00 | 0.74 |
| BD-pTau181/Aβ42 | 37 | 0.75 (0.63-0.87) | 11.02 (2.44–49.71) | 0.00 | 0.76 | 0.74 (0.63-0.86) | 11.02 (2.44–49.71) | 0.00 | 0.76 |
| pTau181/Aβ42 | 37 | 0.64 (0.52-0.77) | 3.03 (0.99–9.26) | 0.04 | 0.64 | 0.64 (0.52-0.77) | 3.03 (0.99–9.26) | 0.04 | 0.64 |
| BD-pTau231/Aβ42 | 37 | 0.61 (0.45-0.76) | 2.00 (0.65–6.11) | 0.21 | 0.59 | 0.60 (0.45-0.76) | 2.00 (0.65–6.11) | 0.21 | 0.59 |
| pTau231/Aβ42 | 37 | 0.68 (0.56-0.79) | 10.64 (1.38–81.83) | 0.00 | 0.70 | 0.67 (0.56-0.79) | 10.64 (1.38–81.83) | 0.00 | 0.70 |
| Aβ40/Aβ42 | 37 | 0.52 (0.33-0.72) | 3.46 (1.13–10.59) | 0.02 | 0.62 | 0.53 (0.33-0.72) | 3.46 (1.13–10.59) | 0.02 | 0.62 |
| Time-dependent discrimination and risk estimates for prediction of amyloid PET progression (A− to A+) among baseline cognitively unimpaired participants. Results are shown for plasma pTau/Aβ42 ratio measures and additional phosphorylated tau species across multiple amyloid positivity thresholds (CL ≥18, ≥24.1, and ≥37). Discrimination is summarized using time-dependent area under the curve (tdAUC) at prespecified 2.5- and 4-year horizons. Hazard ratios (HRs) and C-indices were derived from Cox proportional hazards models comparing high versus low biomarker groups within each horizon. Log-rank p-values compare Kaplan-Meier (KM) curves for the same high versus low biomarker groups. HRs compare high versus low biomarker groups defined by Youden cutoffs within each threshold. In some threshold-specific analyses, estimates were the same at 2.5 and 4 years because most amyloid conversion events occurred before 2.5 years. CL indicates Centiloid. | | | | | | | | | |
